# Effects of COVID-19 infection during pregnancy and neonatal prognosis: what is the evidence?

**DOI:** 10.1101/2020.04.17.20069435

**Authors:** Álvaro Francisco Lopes Sousa, Herica Emilia Félix de Carvalho, Layze Braz de Oliveira, Guilherme Schneider, Emerson Lucas Silva Camargo, Evandro Watanabe, Denise de Andrade, Ana Fátima Carvalho Fernandes, Isabel Amélia Costa Mendes, Inês Fronteira

## Abstract

**Background:** Little is known about how COVID-19 infection affects pregnant women, as well as about the possibility of vertical transmission or complications in childbirth. This study’s aims to assess the current evidence presented in the literature regarding the potential risks of COVID-19 infection among pregnant women and consequent fetal transmission.

**Methods:** a systematic literature review assessing papers published in the most comprehensive databases in the field of health, intended to answer the question: “What are the effects of COVID-19 infection during pregnancy and what is the neonatal prognosis?”

**Results:** 42 papers published in 2020 were eligible. Were included 19 case reports (45%), 15 cross-sectional descriptive studies (35%), 6 cross-sectional analytical studies (14%), one case-control study (3%) and one cohort study (3%), presenting low levels of evidence. A total of 650 pregnant women and 511 infants were assessed. More than half of pregnant women having cesarean deliveries (324/64%). Only 410 (80%) infants were tested for SARS-CoV-2, of which 8 (2%) were positive, however, based on what was assessed that there is no evidence of vertical transmission so far, as there are gaps concerning the care taken during and after delivery, and biological sample proper for testing the SARS-CoV-2.

**Conclusions:** health professionals cannot rule out a possible worsening of the clinical picture of the pregnant woman infected with SARS-CoV-2 because she is asymptomatic or does not have comorbidities related to gestation. Pregnant women and health professionals should be cautious and vigilant, as soon as their pregnancy is confirmed, with or without confirmed infection, as this review checks for infected pregnant women in all trimesters of pregnancy.

## 1. Introduction

The World Health Organization (OMS) declared on January 30th, 2020, that the outbreak of COVID-19, a respiratory disease caused by the new coronavirus SARS-CoV-2, is the sixth public health emergency of international concern. [1-2] Due to this highly transmissible virus, by April 9th, 2020 it had been spread to five continents and approximately 85,522 people had died because of it.[2]

Considering transmission seems to mainly occur through contact with respiratory droplets [3] produced by an infected person, anticipating public health measures intended to control and prevent the infection such as adherence to universal precautions, quarantine, and timely diagnosis are options available to mitigate the transmission of COVID-19.[4]

Clinical manifestations range from asymptomatic cases, mild upper airway infection up to severe and fatal cases with pneumonia, and acute respiratory failure.[5-7] This variation is because people with prior diseases/comorbidities are less apt to fight the virus so that it is more likely to reach the lungs and cause pneumonia. Elderly individuals with comorbidities such as Non-Communicable Diseases and immunocompromised persons are at the highest risk of developing signs and symptoms of COVID-19 and having them worsened. [5-6]

It is, however, unknown how COVID-19 infection behaves in key populations more commonly susceptible to viral diseases, such as pregnant women, [8] as well as whether there is the possibility of vertical transmission to the fetus or premature birth.

The changes in the immune system of pregnant women make them more susceptible to infectious processes, in addition to the manifestations of the infection, with the risk of adverse maternal and neonatal complications, premature birth, spontaneous abortion, application of endotracheal intubation, restriction of intrauterine growth, hospitalization in an intensive care unit, renal failure, intravascular coagulopathy and transmission to the fetus or newborn. [9]

Current studies on the susceptibility of pregnant women to infection by COVID-19 are still incipient and adopt poor methods, and although transmission of the virus to the fetus or baby during delivery or pregnancy has not been proven, the presence of antibodies has already been identified, specific IgG for viruses in neonatal serum samples. [10]

Due to the need to provide evidence for clinical practice involving pregnant women, this study’s objective was to assess current evidence presented in the literature regarding the potential risks of COVID-19 infection among pregnant women and consequent fetal transmission.

## 2. Materials and Methods

This systematic literature review, [11] with no protocol registration, is intended to answer the question: “What are the effects of COVID-19 infection during pregnancy and what is the neonatal prognosis?”. The PECO[12] method was adopted, in which:

- Population (P) = Pregnant women
- Exposure (E)= COVID-19 infection
- Comparison (C)= has not been an object of study
- Outcome (O): Maternal and / or fetal infection by SARS-CoV-2

A search was conducted in the following databases: US National Library of Medicine (PubMed), Scopus, Embase, ScienceDirect (Elsevier), Web of Science (WoS), Scholar Google and preprints servers bioRxiv and medRxiv, as well as the bibliographic references of the selected papers (hand searching). These databases were selected due to their breadth and representativeness in the field of basic and health sciences. Terms that derived from the following expressions were used according to the databases/servers: “COVID-19” OR “SARS-CoV-2” AND “Pregnancy” AND “Perinatal”. To avoid screening biases, two researchers with expertise in the method and topic under study, independently and concomitantly, though in different locations, searched all the databases on May 1^st^ and 2^nd^. The researchers discussed to reach a consensus about which papers would be included or excluded from the study and a third reviewer mediated disagreements that prevented reaching a consensus.

Observational epidemiological studies and case reports addressing the clinical conditions of mother-fetus pairs and including primary data of patients over 18 years old were considered eligible. Manuscripts that contained only data from pregnant women, or only fetuses, or that did not address the period of delivery, such as puerperium, were disregarded. No restrictions regarding the period of publication or language were imposed. Review papers, opinions reports, local reports, abstracts of events, and similar works were excluded. Social, demographic, and clinical data included in the studies were collected.

The GRADE system was used to classify levels of evidence. The results are presented in terms of prevalence calculated in the study and combined results.

## 3. Results

In total, 125 studies were initially identified, 17 of which were excluded because they were repeated. Of the 108 remaining studies, 48 were excluded because they did not address this review’s objective. The full texts of 60 studies were analyzed, though 18 did not meet the eligibility criteria so that 42 studies remained and composed the final sample of this systematic review (Figure 1).

**Figure 1.**
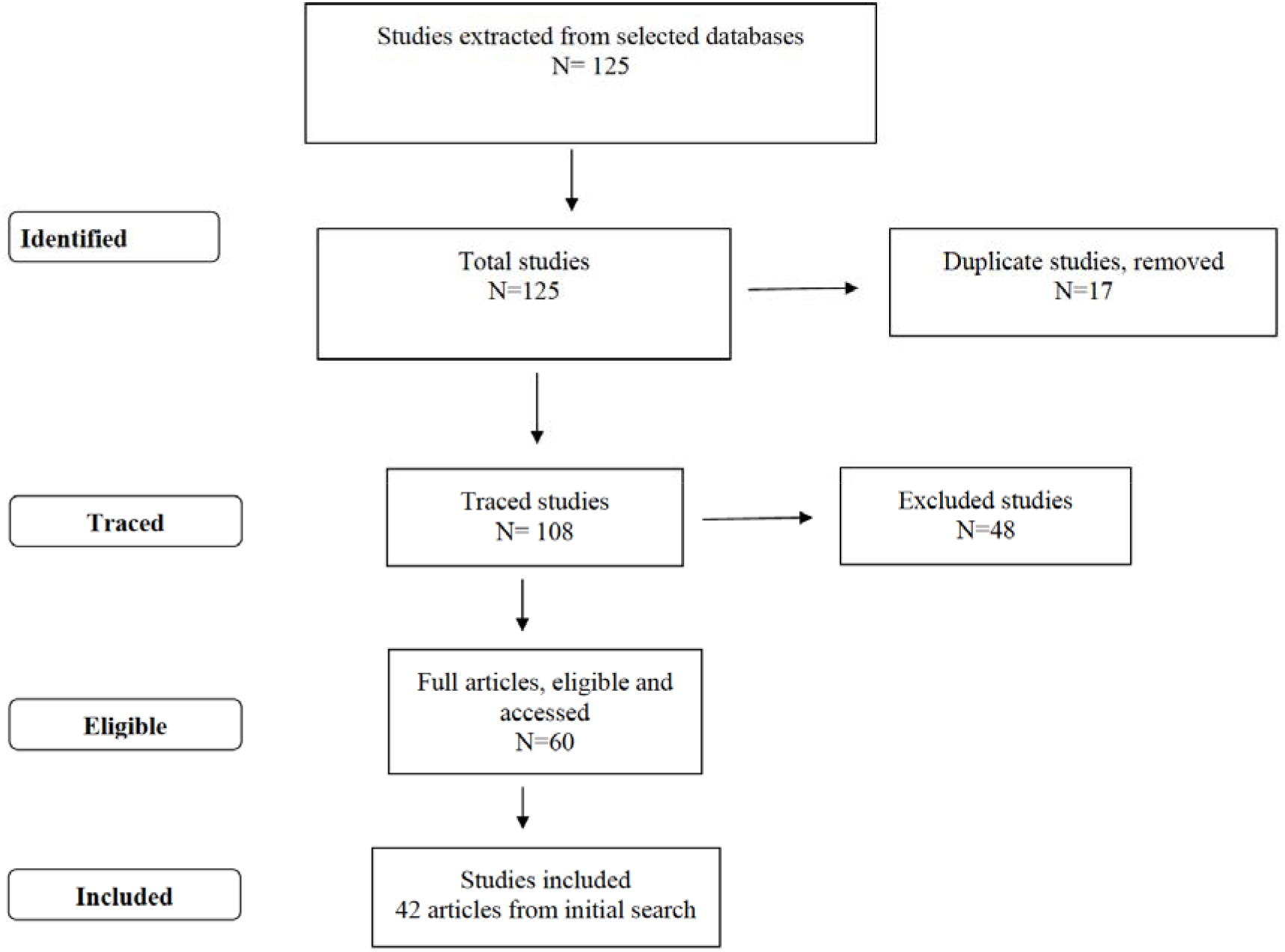
– Study selection flowchart.

The studies were divided into: 19 case reports (45%) [13-31], 15 cross-sectional descriptive studies (35%) [31-46], 6 cross-sectional analytical studies (14%) [47-52], one case-control study (3%)[53] and one cohort study (3%) [54] (Table 01).

**Table 1.**
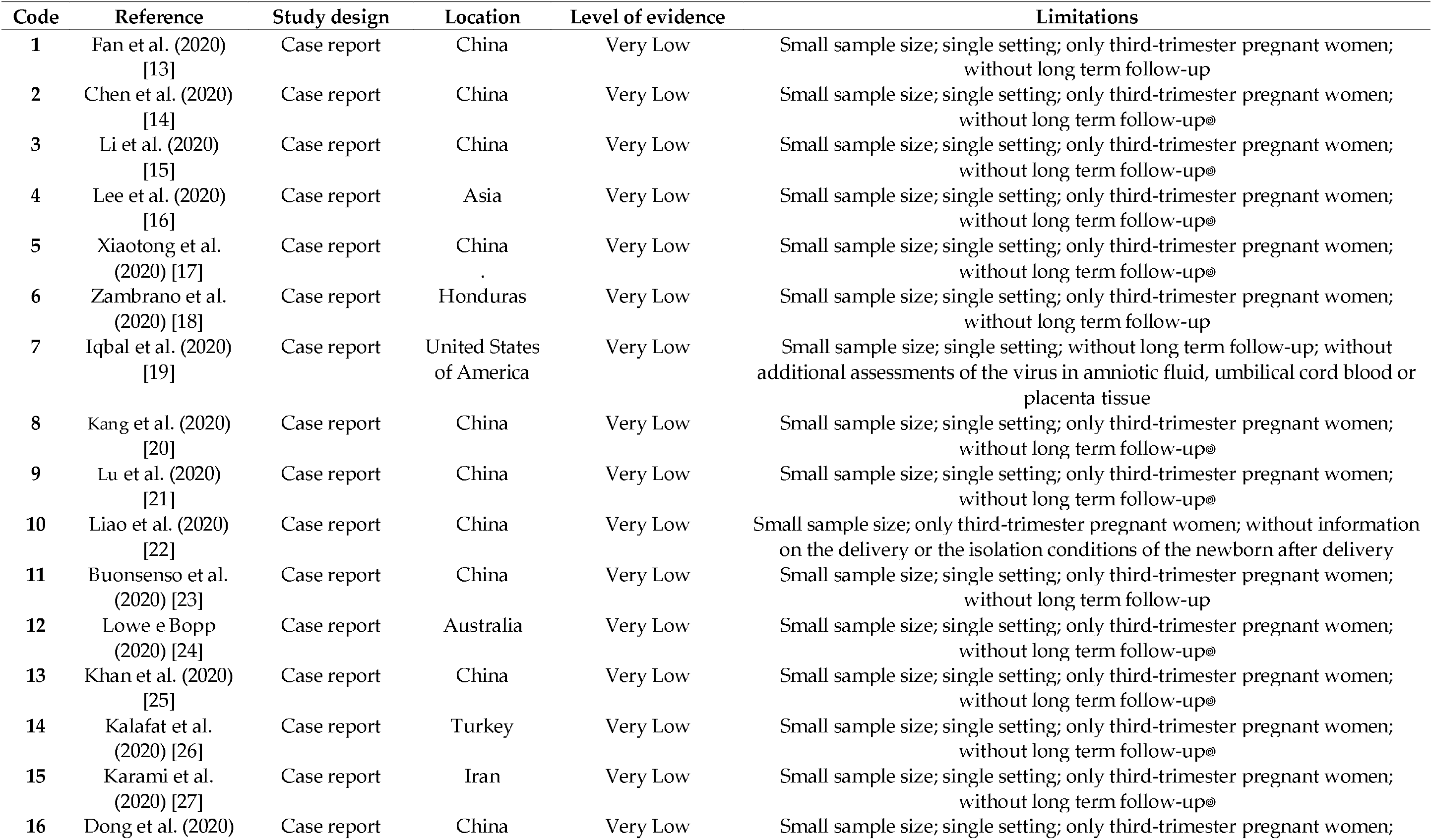
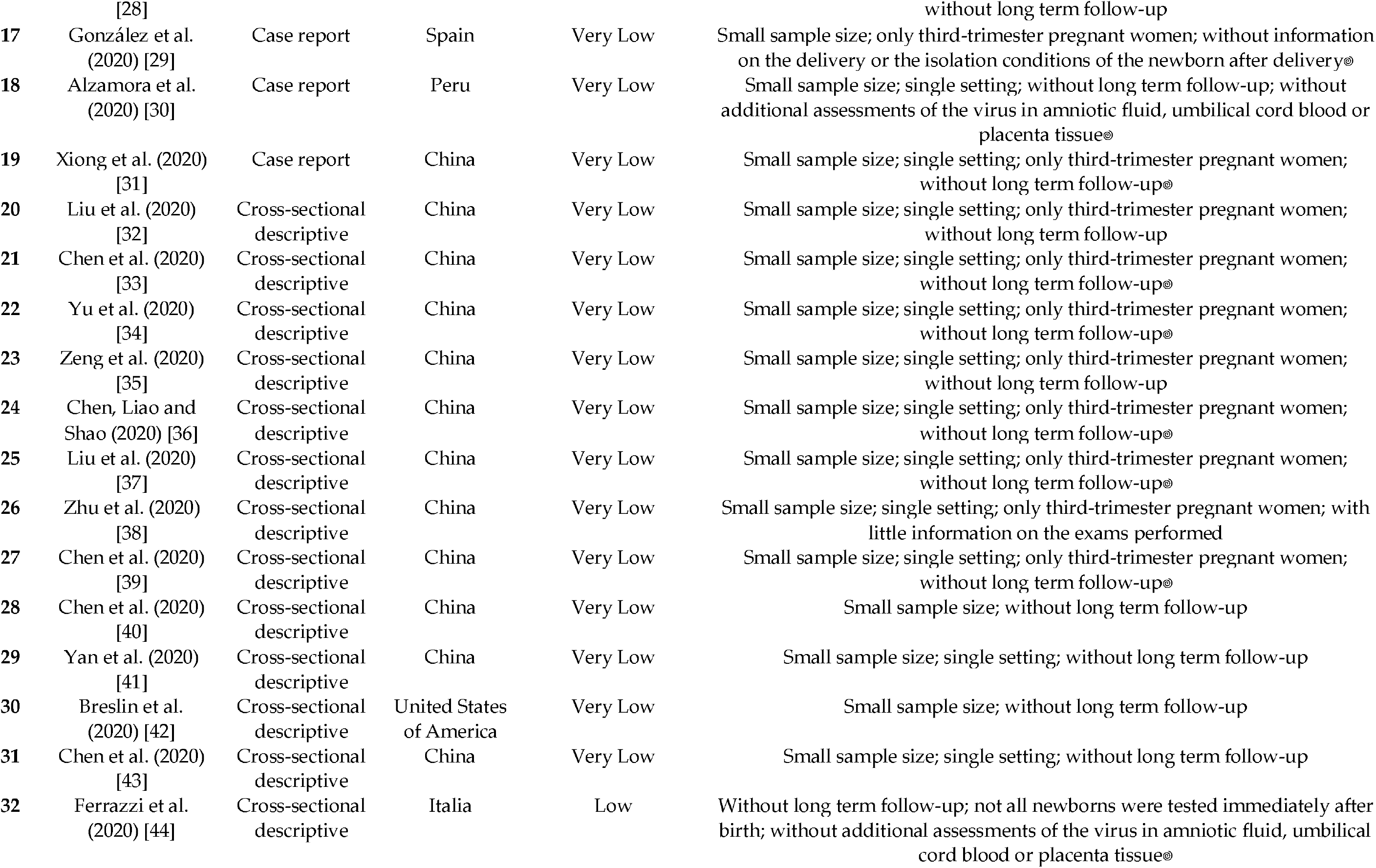
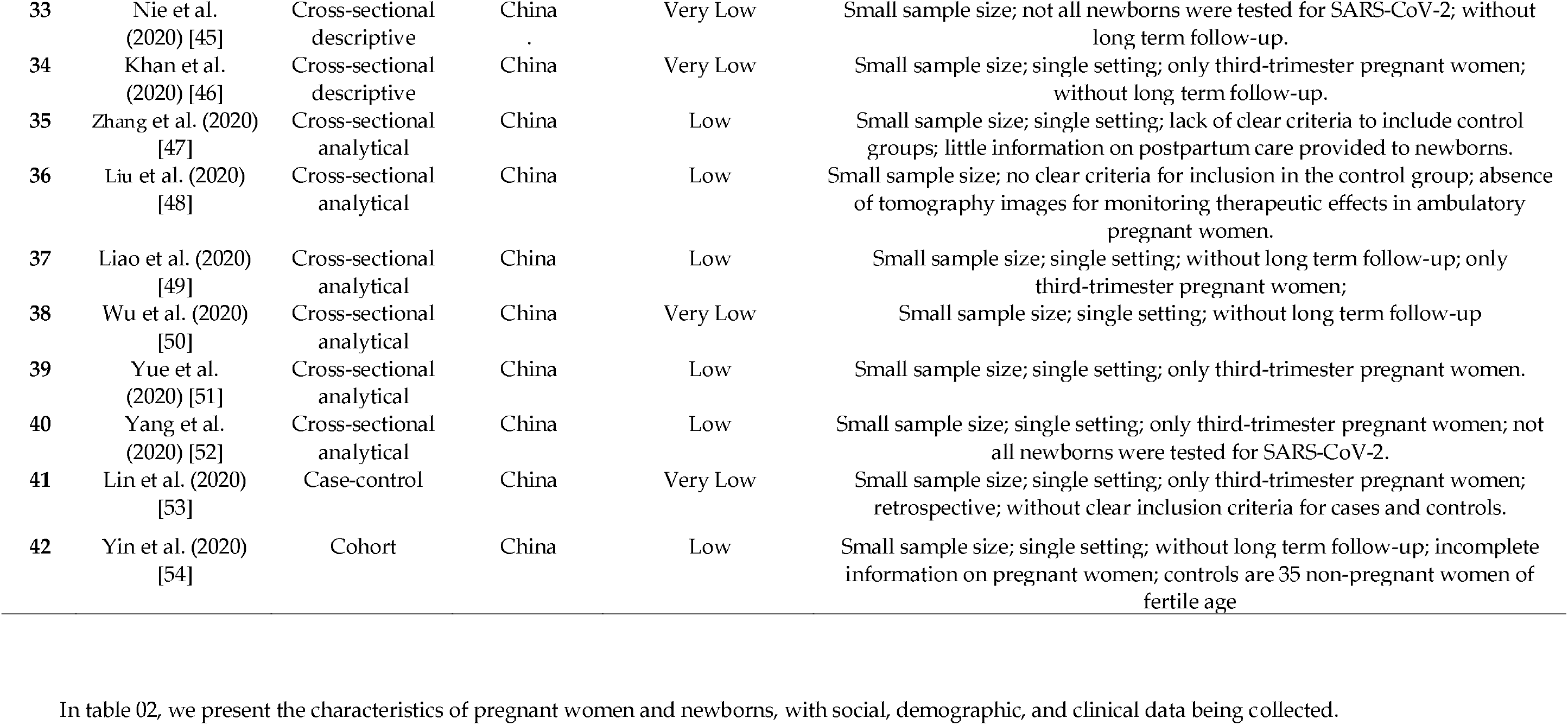
– Characteristics of included studies: reference, study design, location, level of evidence and limitations

In table 02, we present the characteristics of pregnant women and newborns, with social, demographic, and clinical data being collected.

**Table 2.**
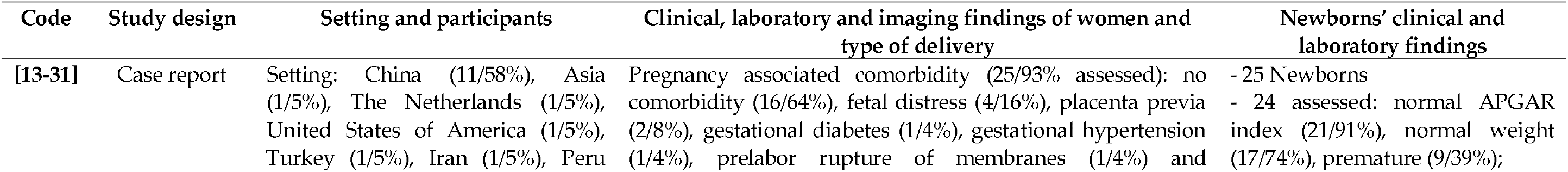
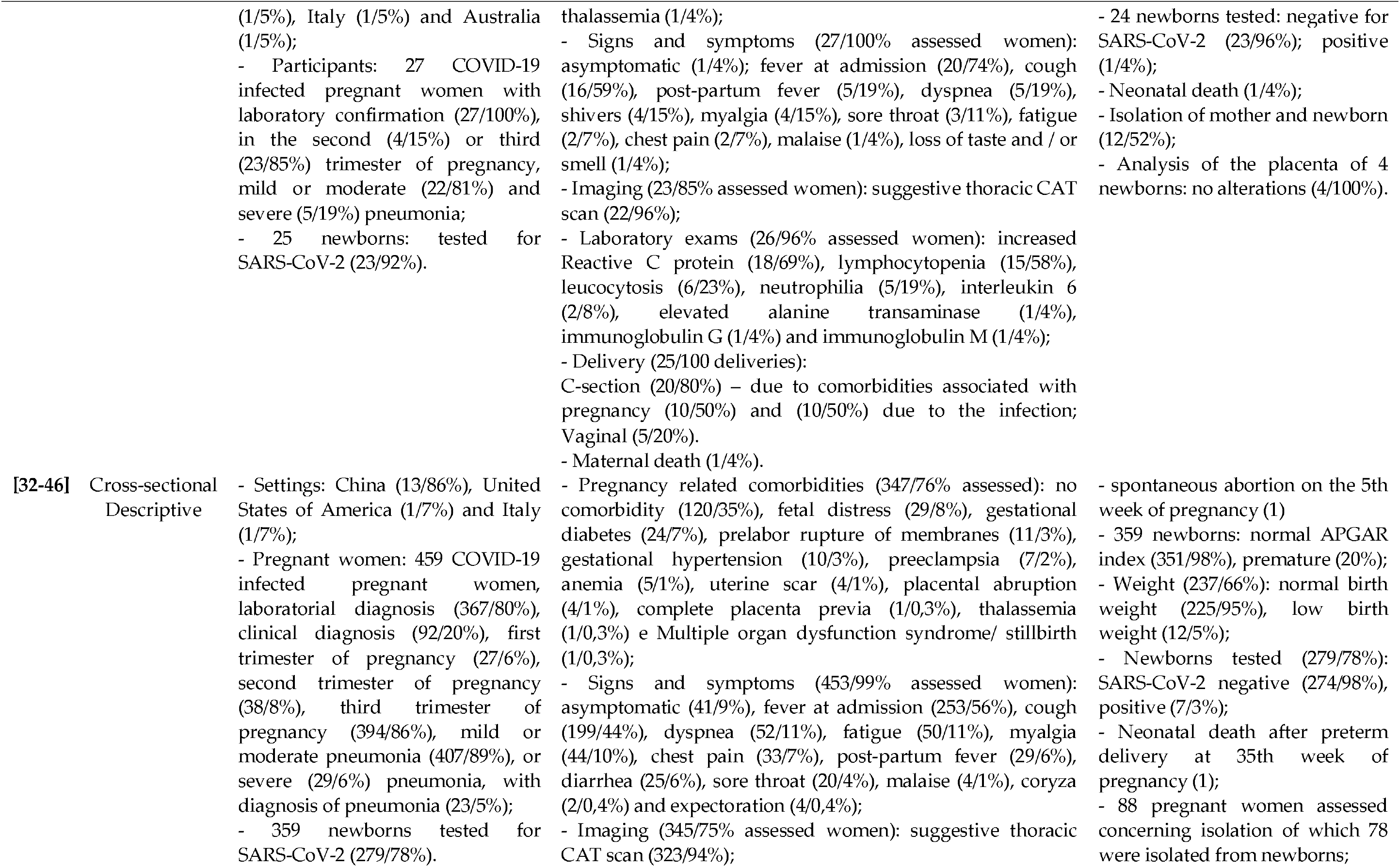
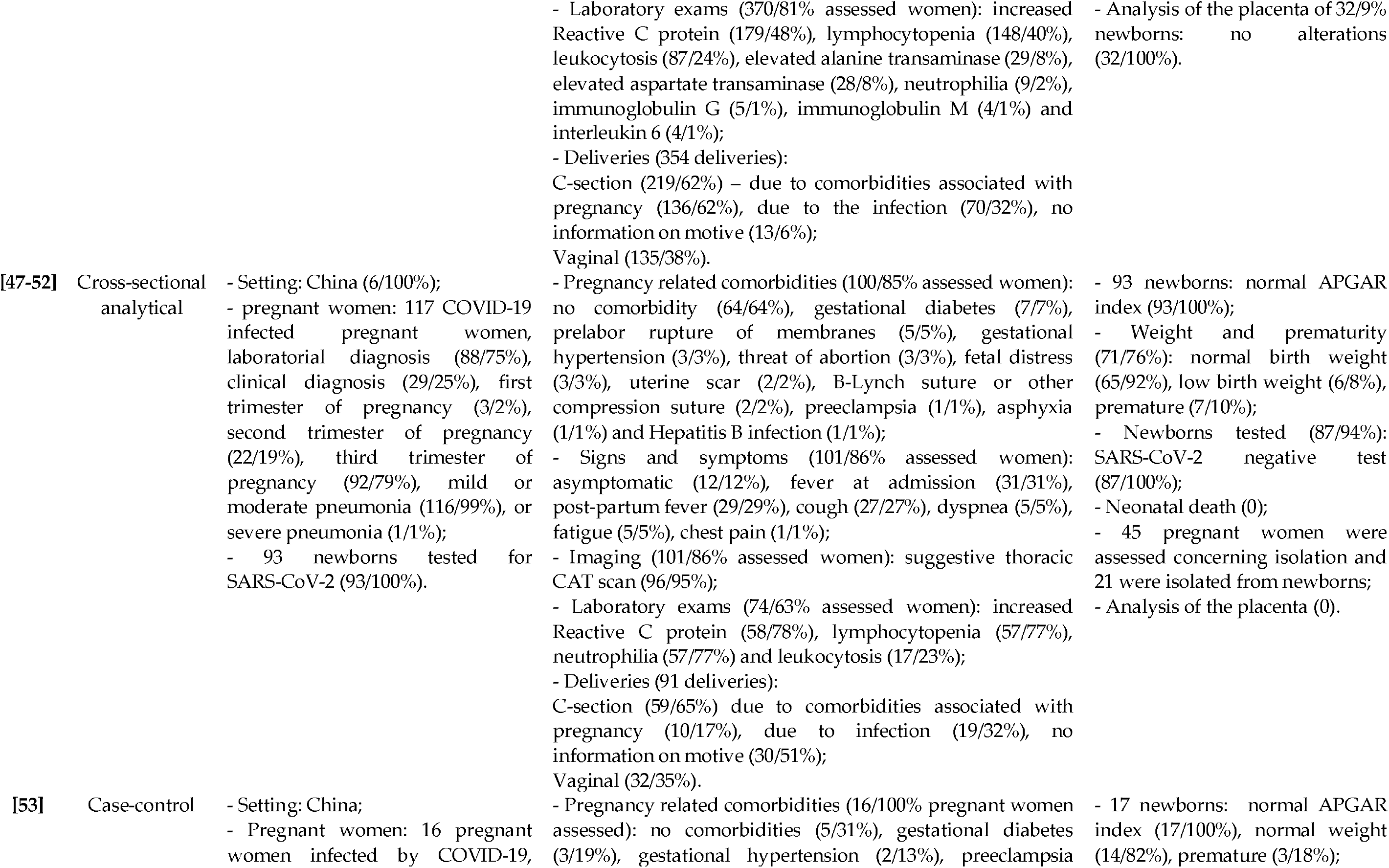
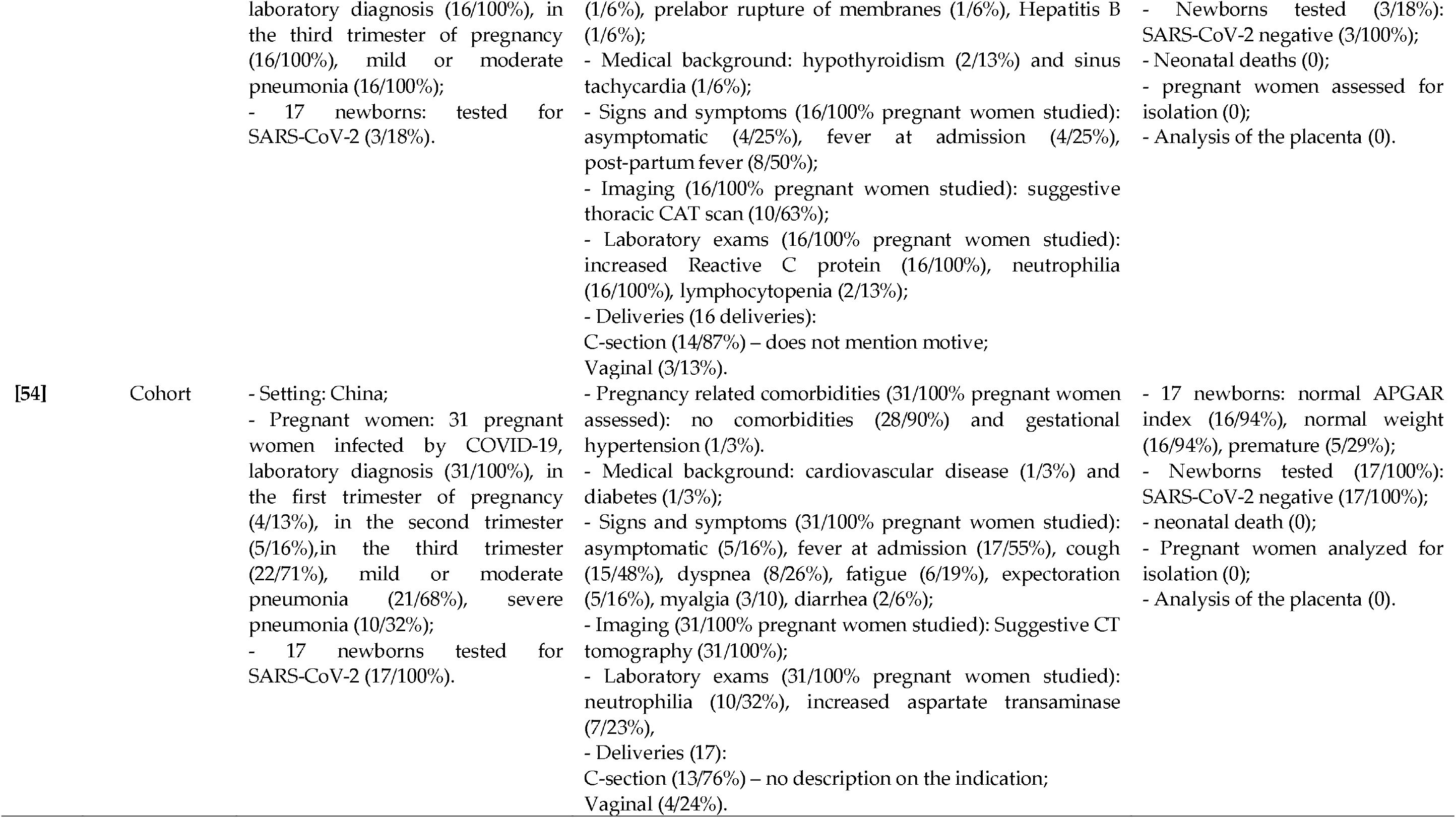
– Characteristics of included studies: study design, setting and participants, clinical, laboratory and imaging findings of women and type of delivery and clinical and laboratory findings of newborns

The 19 case reports (45%)[13-31] addressed 27 women in the second (15%) and third trimester (85%) of pregnancy, admitted with signs and symptoms of COVID-19 (96%), later confirmed through RT-PCR (100%). Twenty-two (81%) developed mild to moderate pneumonia. The majority of the studies were conducted in China (58%) [13-15,17,20-23,28,31] and were limited to the description of the main features of the disease in positive pregnant women. As for comorbidities, more than half of the women did not present any comorbidity(64%)[13,17,19,21-25,27-29,31]. Among those with comorbidities, the most common one was fetal distress (16%)[16,20,22-23]. The most clinical signs and symptoms were fever (74%)[13-14,16,19,22-31] and cough (59%) [15-16,19-23,25-27,29,31]. Among the imaging findings, chest tomography revealed pulmonary changes (96%) [13-17,19-23,25-27,29-31]. Laboratory exams revealed increased reactive C protein (69%)[14-15,17,19,20,23,25,27,29-31] and decrease in lymphocytes (lymphocytopenia) (58%) [13-14,17,19-20,22-23,26-29,31]. Twenty-five newborns were born from twenty-five women, predominately through C-section delivery (80%)[13-17,20-23,25-26,28-30] for half of which there was medical indication due to maternal comorbidities [14-16,20,22-23,26,30] or due to the infection [13,17,20,23,25,28-29]. Of the 25 newborns, one died [27] along with his/her mother, and 24 were tested for SARS-CoV-2: one was positive [30]. In half of the cases, newborns were isolated from mothers (52%)[13-14,16-17,19-21,28,30] and in four cases the placenta was analyzed for pathological alterations [13,17], There were no cases of vertical transmission.

In cross-sectional descriptive studies (35%)[31-46], 11 (86%) were conducted in China [32-41,45-46]. A total of 459 pregnant women were assessed for COVID-19, of which 367 (80%) were diagnosed with the disease through RT-PCR. The majority (86%) were in the third trimester of pregnancy and had mild to moderate pneumonia (89%). The most frequent were women not to present any comorbidity (35%) [32-34,36-37,39,41,43-46]. The signs and symptoms more frequently found in pregnant women were fever [32-34,37-45] and cough [32-34,36-46]. Imaging findings revealed suggestive images of infection in 323 (94%) pregnant women [33,41,43,45-46] and laboratory exams showed increased reactive C-protein (48%) [33-34,36-37,39-41,43-44] and lymphocytopenia (40%)[33-34,36-37,39-41,43-44], 354 pregnancies resulted in 359 newborns with 5 twin births. C-section was the most frequent type of delivery (62%), the majority resulting from pregnancy comorbidities (62%) [32-34,36-42,44-46]. A newborn died after birth [32] and there was a stillbirth [41]. Two hundred and seventy-nine newborns were tested for SARS-CoV-2, of which 7 tested positive (3%) [34,44-46]. Isolation of the newborn from the mother occurred in 78 (87%) [42,44-45] cases. A total of 32 placentas were analyzed with no abnormal findings [45].

All cross-sectional analytical studies (14%) [47-52] were conducted in China, in a total of 117 COVID-19 infected pregnant women, with laboratory (75%) and clinical diagnosis (25%). Most pregnant women were in their third trimester (79%) and had developed mild to moderate pneumonia (99%). Clinical findings showed that only 36 (36%) [48-51] women had some type of comorbidity (Table 2). The most common signs and symptoms were fever at admission [48-52], post-partum fever [48,51-52] and cough [48-50,52].

Chest computed tomography was suggestive in 96 (95%) women [48-52] and laboratory exams revealed increased reactive C protein (78%), [48-50], lymphocytopenia (77%)[48-50] and neutrophilia (77%) [48,50], Of the 91 women who delivered, 59 (65%) had a C-section of which 19 were due to COVID-19 [47-,51], A total of 93 babies were born with one set of twins. Eighty-seven (94%) were tested for SARS-CoV-2 and all were negative. Twenty-one (47%) pregnant women were isolated from their newborns [49,52], There were no neonatal deaths, no placentas were analyzed and there was no confirmation of vertical transmission.

The case-control study (3%)[53] was conducted using the medical records of pregnant women admitted to a hospital in China. The study compares the clinical features, maternal and neonatal outcomes of 16 pregnant women with COVID-19 and 18 without the disease but suspected. There are no clear definitions for the inclusion of participants in the case or the control group. The description in Table 2 relates to COVID-19 infected pregnant women. All 16 women were in their third trimester of pregnancy, of which 5 (31%) had no pregnancy-related comorbidity. The signs and symptoms more frequent were fever at admission (25%) and after birth (50%). Ten women had suggestive tomography and there was an increase in reactive C protein and neutrophilia in all studied women. Fourteen (87%) women underwent C-section delivery. The study does not mention what was the indication for having this type of delivery. A total of 17 babies were born with one set of twins, with no complications. Only three newborns were tested for SARS-CoV-2 and were all negative. There was no description of isolation measures after birth or analysis of the placenta.

The cohort study (3%)[54], also conducted in China, retrospectively describes 31 pregnant women and 35 non-pregnant women with COVID-19. Only the clinical findings of pregnant women are described. In total 31 pregnant women were assessed. The majority was in the third trimester of pregnancy (71%), all had confirmation of diagnosis through RT-PCR, 21 (68%) developed mild to moderate pneumonia, and 10 severe pneumonia. Twenty-eight did not present comorbidities during pregnancy. The most prevalent signs and symptoms were fever (55%) and cough (48%). Chest tomography was suggestive in all cases and abnormal laboratory tests were related to the increased number of neutrophils (32%), aspartate transaminase (26%), and interleukin 6. There were 17 deliveries were performed, 13 (76%) of which were C-sections, the authors do not describe the reason for the indication. 17 babies, single, healthy fetuses were born and tested negative for SARS-CoV-2. There was no description of the isolation of the mother and baby after delivery and no evaluation of the placentas.

## 4. Discussion

This review was intended to answer a question concerning the effects of COVID-19 infection during pregnancy and neonatal prognosis. Forty-two studies were eligible and included case reports, cross-sectional, analytical cross-section, case-control, and cohort presenting low levels of evidence. The low levels of evidence are due to the novelty of COVID-19 pandemic and the need for rapidly acquiring knowledge to support public policies. As the number of cases increases worldwide, evidence about the impact of this virus during pregnancy for both women and newborns is expected to become stronger, especially with the development of more robust comparative studies and follow-up with control groups.

A total of 650 pregnant women were assessed, 557 were from China, [13-15,17,20-23,25,28,31-41,43,45-54] 44 EUA, [19,42] 42 Italy, [44], and 1 and 1 pregnant woman in Asia,[16] Honduras,[18] Australia,[24] Turkey,[26] Iran,[27] Spain [29] and Peru.[30] All women evaluated were in the fertile period and, as for the time of pregnancy, there were pregnant women in the first trimester (34/5%), in the second (69/11%) and third trimester of pregnancy (547/84%), explaining why only 511 infants were born in the period. The fact that most pregnant women were from China imposes a considerable challenge to interpret evidence, considering cultural and epidemiological differences when comparing with pregnant women from other countries and cultures. However, even in a minority (93/14%), non-Chinese pregnant women were evaluated and the characteristics (clinical and epidemiological) showed no differences.

Regarding the pregnant women’s age, the fertile period, and the length of pregnancy, the studies analyzed showed a wide variation and a lack of evidence of infection by SARS-CoV-2 during the first and second trimester of pregnancy. It can be inferred that, according to the low prevalence of severe infection among pregnant women (45/10%), many of them could be asymptomatic and/or with mild symptoms, without the need for hospital care. Corroborating this data, a Norwegian cohort study of 1258 pregnant women during the influenza pandemic in 2009 showed that 226 (18%) had influenza (H1N1) and only three were hospitalized. It is noteworthy that most pregnant women were in the first or second trimester of pregnancy. The study authors state that there is little evidence that mild influenza during pregnancy is associated with an increased risk of preeclampsia as well as premature and small birth-weight babies for gestational age [55].

Regarding the Middle East Respiratory Syndrome (MERS), there are limited data on the prevalence and clinical characteristics of MERS during pregnancy, birth, and the postnatal period. A systematic review with meta-analysis recovered seven studies, which did not report spontaneous abortion. The rate of premature birth was 32.1% (3 of 11), all occurring before 34 weeks of gestation. Preeclampsia was described in 19.1% (1 of 7), however, no cases of premature rupture of membranes or restricted fetal growth were reported. The rates of cesarean delivery and perinatal death were 61.8% (5 of 8) and 33.2% (3 of 10), including two stillborn and one neonatal death (4 hours after the birth of an extremely premature baby), respectively. There were no reports of fetal distress, Apgar score <7 at 5 minutes, neonatal asphyxia, and admission to the neonatal intensive care unit (ICU). Finally, signs of vertical transmission were not found during the follow-up period in any of the newborns [56].

According to the above, it appears that the limited data on infection with the new coronavirus in early pregnancy may be related to the absence of tests performed during this period, as Apgar there are asymptomatic pregnant women, with mild signs and symptoms and there is no screening of these pregnant women since the beginning of pregnancy, this infection may go unnoticed or be detected only after delivery. Thus, we suggest that tests for COVID-19 should be performed as a routine in prenatal care.

It is important to note that the fact that the majority of the women evaluated in this study are at the end of pregnancy, does not mean that there is a higher prevalence of cases in pregnant women in the third trimester, because, as we have already discussed, there is not enough data in the scientific literature such as, for example, monitoring pregnant women from the beginning of pregnancy until delivery. Thus, if we calculate the time of a normal pregnancy and buy the period that the first case of COVID-19 (December 2019) was described, this pregnant woman would not have given birth yet.

About diagnosis, the present study verifies that of the 650 pregnant women evaluated, 529 (81%) and 121 (10%) were diagnosed by reverse transcription followed by polymerase chain reaction (RT-PCR) and clinically, respectively. The gold standard for diagnosing Covid-19 is tissue culture in which the antigen is isolated, using polymerase chain reaction (PCR), which detects nucleic acid. Even so, a single result not detected through RT-PCR for SARS-CoV-2 does not exclude a COVID-19 diagnosis, as there are various factors as e.g., inadequate sample collection, type of biological sample, the time elapsed between sample collection and onset of symptoms, fluctuation of viral load, that may influence a test’s result. For this reason, an RT-PCR test should be repeated in another sample of a patient’s respiratory tract whenever there are discordances between results and epidemiological conditions, especially in populations where a false positive may result in disastrous consequences.

In total, [13-54] comorbidities related to pregnancies were evaluated and described in 514 (79%) pregnant women, of which 234 (46%) did not present any comorbidity. On the other hand, pregnant women who presented one or more comfort, prevailed, fetal distress (36 cases), gestational diabetes (34 cases), gestational hypertension (17 cases), and premature rupture of the membranes (17 cases). It is worth noting that the studies only reported the number of pregnant women with comorbidities, however without describing whether one or more comorbidities.

The presence of comorbidities related to pregnancy does not seem to directly influence the adverse outcomes of pregnant women and their newborns, as the two neonatal deaths were of mothers without comorbidities, but who for some reason developed severe pneumonia. However, it is observed that fetal distress was the most prevalent comorbidity, demonstrating that the conditions of the fetus should be carefully evaluated, especially, in those asymptomatic and without comorbidities. Thus, the lack of comorbidity can influence the care and attention given to pregnant women so that health professionals can discriminate against detailed evaluations. [57]

Concerning the signs and symptoms of infection at the time of admission, 626 (96%) pregnant women were evaluated[13-54], and the main signs and symptoms presented were fever at admission (325/56%), coughing (273 / 44%) and dyspnea (70/11%). It is remarkable that 63 (10%) of pregnant women were asymptomatic but were tested (through RT-PCR to detect SARS-CoV-2) due to exposure to people diagnosed with COVID-19. They tested positive and were hospitalized. Such a fact reinforces the need to follow recommendations provided by the Centers for Disease Control and Prevention (CDC) to testing risk groups in contact with those diagnosed with COVID-19, [58] though this approach may not be feasible in some contexts where there is a shortage of tests.

Regarding imaging tests, chest computed tomography (CT) was performed in 516 (79%) pregnant women [13-54], with 482 pregnant women showing changes suggestive of infection[13-17,19-23,25-27,29-31, 33.41.43.54-46.48-52.53-54], CT was very useful in the initial assessment at the time of admission. The most prevalent changes were in the findings of C-reactive protein, which was above the normal range[14-15,17,19,20,23,25,27,29-31,33-34,36-37,39-41,43-44,48-50,53] and lymphocytopenia [13-14,17,19-20,22-23,26-29,31,33-34,36-37,39-41,43-44, 48-50], Also, other tests to a lesser extent showed changes: neutrophilia, leukocytosis, increased alanine transaminase, aspartate transaminase, interleukin 6, immunoglobulin G, and immunoglobulin M. It is outstanding that these changes in CT and laboratory findings are found in studies with the general population. [59] Concerning the evaluation of imaging tests, whether they are chest X-rays or chest CTs, these can assist in the diagnosis of the disease, however, they should not be taken as unanimity for confirmation or exclusion of SARS-CoV-2 infection. This can be explained by the fact that different bacterial and viral etiologic agents cause pulmonary infections. In this way, imaging tests such as chest CT, widely used during emerging respiratory outbreaks or as in the case of the pandemic currently established by COVID-19, have high sensitivity, but low specificity. Despite the limitations, these tests should be used to screen, evaluate, and monitor this kind of infection. [60] 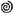

Concerning childbirth, 503 were reported, with more than half of pregnant women having cesarean deliveries (324/64%). When checking the indications for cesarean section, in 156 pregnant women, pregnancy-related comorbidities were the cause, 99 were described in the studies as an indication for infection, and 69 did not provide a description of the indication. Regarding the indication for cesarean section because of the infection, the indications were not reported. In most cases, the time of delivery was determined by obstetric indications, rather than the maternal diagnosis of COVID-19.[61]

The decision for the type of delivery is usually more influenced by the presence of maternal and/or fetal impairment. When there were imminent risks, emergency cesarean section was the alternative, which has happened in the case of SARS-CoV-2 infections in which the pregnant woman’s clinical condition is complex. However, in the presence of COVID-19, the threshold for cesarean delivery became lower than usual so that infection control procedures could be more easily adhered to and the transmission of the disease to the fetus minimized. [62]

Regarding the characteristics of the newborns, 511 babies were born, being 503 single, and 8 twins. The normal Apgar score and normal birth weight were verified in 498 (97%) and 337 (66%) newborns, respectively. A total of 410 (80%), 96 (19%) and 10 (2%) newborns were tested for SARS-CoV-2 [8 (2%) were positives], premature and underweight at birth (<2,500g), respectively [30,34,44-46]. Two neonatal deaths [27,32] and one spontaneous abortion [41] have been reported. After delivery, isolation between mothers and newborns was carried out in 117 pregnant women [13-14,16-17,19-21,28,30,42,44-45], as well as no changes were found placental in the 36 samples evaluated [13,17,45].

It is observed that the majority of newborns did not have serious complications. The unfavorable outcomes refer to two neonatal deaths, a spontaneous abortion, a maternal death, together with a stillbirth, eight (2%) positive SARS-CoV-2 tests, and three newborns with high rates of IgG and IgM antibodies against SARS-CoV-2.

Concerning neonatal deaths, the first death was described in the study by Liu et al. [32], a 31-year-old pregnant woman in her third trimester of pregnancy was confirmed laboratory with SARS-CoV-2. Although the pregnant woman had no previous illnesses, she had complications and was admitted to the ICU due to Acute Respiratory Distress Syndrome (ARDS), Acute Liver Failure, Acute Kidney Failure, Multiple Organ Dysfunction Syndrome (MODS) and septic shock. The pregnant woman was intubated, received mechanical ventilation, and an emergency cesarean section was performed due to premature labor. The fetus was born with an Apgar score of 1 and did not survive. The mother recovered. According to the study, the serological evidence was not suggestive of vertical transmission of the virus.

The second case is described by Yan et al.[41], a 32-year-old pregnant woman was confirmed with COVID-19 by RT-PCR in the third trimester of pregnancy. After the hospital admission, her condition worsened, she developed severe pneumonia and septic shock, requiring ICU admission for endotracheal intubation and invasive ventilation. She underwent a cesarean section in her 35th gestational week and gave birth to a newborn baby who remained with Apgar of 1 in the 1st, 5th, and 10th minutes after delivery. Severe neonatal asphyxia and the need for invasive ventilation have been reported, but the newborn died within two hours of birth. In this same retrospective study, the authors briefly cited the case of a woman, in the 5th gestational week, who had a spontaneous abortion. It is believed that the brevity of the data description was due to the lack of information in the medical records.

Karami et al.[27] reported the death of a mother and her baby. The pregnant woman was 27 years old, in the 30th gestational week, and SARS-CoV-2 infection diagnosed by RT-PCR only after delivery. The patient had no underlying disease, however, she had fever, cough, myalgia, and dyspnea and was referred to the hospital. Due to tachypnea, oxygen saturation below 95%, tachycardia, and high fever, the patient had to be transferred to the ICU. Abnormal findings were found in imaging tests, leukopenia, thrombocytopenia, elevated levels of C-reactive protein, and lactate dehydrogenase. Due to a worsening of clinical symptoms, the patient required endotracheal intubation and invasive ventilation. The following day, premature spontaneous delivery occurred vaginally to a fetus with an Apgar score of 0 during the 1st and 5th minute after birth, which did not change with the attempt at neonatal cardiopulmonary resuscitation. After new atypical laboratory and imaging tests, the medical team became suspicious of acute autoimmune vascular disease, initiating treatment without success. The mother died of multiple organ failure.

Regarding the eight newborns who tested positive for SARS-CoV-2, two of them tested positive [46], immediately, after delivery, however they tested negative after 24 hours of delivery. Another newborn tested positive 36 hours after delivery [34], It is important to note that, in these three cases, the isolation or not of the mother and babies after delivery is not described. Besides the authors of these three studies assumed the lack of evidence for vertical transmission due to the absence of intrauterine tissue and amniotic fluid samples.

Two newborns tested positive for the new coronavirus after being breastfed by their mothers without wearing masks, as the maternal infection was not known in the postpartum period. However, vertical transmission could not be confirmed or ruled out, as the authors stressed that testing for COVID-19 was not performed on newborns, immediately after birth. Moreover, in this same study, a pregnant woman with COVID-19 gave birth to a newborn by vaginal delivery, who tested positive for SARS-CoV-2, despite the mother’s wearing a surgical mask and the medical team wearing the adequate PPE throughout labor [44], The other two newborns tested positive 16 hours[30] and 53 hours[45] after delivery, respectively. Both studies described that the guidelines were followed: the use of a mask by the mother, the entire medical team was attired, the mother and baby were isolated after delivery and there was no breastfeeding. However, the authors revealed an important limitation of the study, the absence of complementary assessments (presence of viruses in amniotic fluid, umbilical cord blood, or placental tissue). Of the eight positive babies, two were intubated[30,44], three had mild pneumonia[34,45-46], but in a few days, they were fully recovered.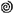

Dong et al.[28] reported the case of a newborn who, shortly after birth, had leukocytosis, a high rate of IgG and IgM antibodies against SARS-CoV-2 and IL-6 cytokines, however, the baby had no symptoms and the test for the virus it was negative. Although the newborn was tested negative for SARS-CoV-2, the authors stated that the high rate of IgM antibodies within 2 hours after birth suggested the occurrence of intrauterine infection, as there would be no transfer of these antibodies from the mother to the fetus through the placenta due to the size of this macromolecule and, in general, they take three to seven days to be produced by the body after contact with the infectious agent, however, they do not rule out the possibility of infection during delivery. 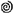

Zeng et al. [35] described similar results regarding the assessment of the presence of specific antibodies against SARS-CoV-2 in the blood of newborns from mothers with confirmed COVID-19 infection. Two babies had rates of IgG and IgM antibodies specific to the virus above the normal level, but none showed symptoms of the infection. The authors of this study stressed the possibility that the newborn developed IgM antibodies during the gestational period if the virus had crossed the placental barrier. 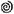

Finally, the studies recovered in this review have several limitations, among them, the fact of the reduced sample size; use of a single institution; retrospective evaluation of medical records where incomplete data exist and this can be observed by checking the number of pregnant women who were recruited and those who were evaluated for comorbidities, signs and symptoms, tests, as well as the babies who were born and those who were tested for infection; the use of the database of notification systems where different authors may have included the same pregnant women, however, this fact may have been positive if the justification is to carry out a different approach and increase the sample size for possible comparisons; most of the cases described are in Chinese pregnant women; absence of a standard of assessment in newborns such as the presence of viruses in amniotic fluid, umbilical cord blood or placental tissue; scarce information about care during delivery and postpartum; absence of clear criteria for inclusion in a control group. However, even with these limitations, the strengths of this review is the ability to compare all the literature available so far, compile loose data and group it more clearly for analysis and inferences and, state, based on what was assessed that there is no evidence of vertical transmission so far, as there are gaps concerning the care taken during and after delivery, and biological sample proper for testing the SARS-CoV-2.

## 5. Conclusions

In summary, the results found in this review show that health professionals cannot rule out a possible worsening of the clinical picture of the pregnant woman infected with SARS-CoV-2 because she is asymptomatic or does not have comorbidities related to gestation. Pregnant women and health professionals should be cautious and vigilant, as soon as their pregnancy is confirmed, with or without confirmed infection, as this review checks for infected pregnant women in all trimesters of pregnancy. Thus, in times of this pandemic, it is important to monitor and screen pregnant women for COVID-19 from the time pregnancy is confirmed.

## Data Availability

Available upon request to the authors

## Author Contributions

AFLS, HEFC, LBO, GS, and IF had the idea for and designed the study and had full access to all of the data in the study and take responsibility for the integrity of the data and the accuracy of the data analysis. AFLS, HEFC, LBO, GS, IF, ELSC, EW, DA, AFC, and IACM did the analysis, and all authors critically revised the manuscript for important intellectual content. All authors gave final approval for the version to be published. All authors agree to be accountable for all aspects of the work in ensuring that questions related to the accuracy or integrity of any part of the work are appropriately investigated and resolved.

